# Impulse Control Disorders Are Independent of Parkinson Disease Motor Subtypes

**DOI:** 10.64898/2026.09.04.26362266

**Authors:** M. Eymaz Chhotani, William Saban, Malco Rossi, Sheng-Han Kuo, Leila Montaser-Kouhsari, Chi-Ying R. Lin, the Parkinson’s Precision Medicine Initiative

**Affiliations:** Department of Neurology, Baylor College of Medicine, Houston, TX USA; Department of Occupational Therapy, Gray Faculty of Medical & Health Sciences, Sagol School of Neuroscience, Center for Accessible Neuropsychology, Tel Aviv University, Tel Aviv, Israel; Sección de Movimientos Anormales, Departamento de Neurología, Fleni, Buenos Aires, Argentina; Instituto Fleni-CONICET (INEU), Buenos Aires, Argentina; Department of Neurology, Columbia University Irving Medical Center, New York, NY USA; Department of Neurology, Brigham and Women’s Hospital, Harvard University, Boston, MA, USA

## Abstract

**Objective:** Impulse control disorders (ICDs) are clinically important nonmotor features of Parkinson’s disease (PD). Tremor-dominant PD (TD-PD) has been associated with greater cerebello-thalamo-cortical involvement, whereas postural instability/gait difficulty-dominant PD (PIGD-PD) has been linked to more prominent basal ganglia and frontostriatal dysfunction. We therefore examined whether ICD symptoms differ by motor phenotypes.

**Methods:** This cross-sectional study analyzed baseline data from the Parkinson’s Precision Medicine Initiative (PPMI). Motor subtype classification was based on Movement Disorder Society-Unified Parkinson’s Disease Rating Scale (MDS-UPDRS) criteria. ICD symptoms were assessed using the Questionnaire for Impulsive-Compulsive Disorders in Parkinson’s Disease-Current Short Form (QUIP-CS) and evaluated across multiple levels of analysis using the standard screening definition and normalized QUIP-CS scores analyzed categorically and continuously. Temporal-alignment, subgroup, multivariable, and Bayesian analyses were performed to assess the robustness of the primary findings.

**Results:** We analyzed 305 participants (173 TD-PD, 132 PIGD-PD). Screening-defined ICD positivity was similar between subtypes (21.4% vs 21.2%, OR = 1.01, 95% CI 0.58-1.76, *p* = 0.970). Continuous and categorical normalized QUIP-CS scores likewise did not differ between groups (continuous score: 0.0338 vs 0.0384, *p* = 0.674; categorical score: 3.5% vs 4.5%, *p* = 0.632). Temporally aligned, subgroup, and multivariable sensitivity analyses consistently supported the primary finding, and Bayesian analysis provided moderate evidence supporting the null hypothesis (BF_01_ = 7.21).

**Conclusions:** ICD symptoms do not differ between TD-PD and PIGD-PD motor subtypes in early PD. These findings suggest that motor subtype-associated differences in cerebellar involvement do not substantially influence ICD symptoms in early PD.

## INTRODUCTION

Parkinson’s disease (PD) is a heterogeneous neurodegenerative disorder characterized by a broad spectrum of motor and non-motor manifestations. Although cardinal motor features define the clinical diagnosis, non-motor symptoms increasingly are recognized as major contributors to functional impairment and reduced quality of life. Among these, impulse control disorders (ICDs) in PD encompass maladaptive reward-related behaviors that substantially affect patient safety, psychosocial functioning, and disease management.^1–3^ These behaviors are frequently associated with dopaminergic therapy but may also reflect disease-related vulnerabilities in cognitive control and reward processing.^3^ The Questionnaire for Impulsive-Compulsive Disorders in Parkinson’s Disease-Current Short Form (QUIP-CS) is a validated screening instrument for assessing ICD symptoms.^4^

In parallel with non-motor heterogeneity, PD demonstrates marked motor phenotypic variability. Patients are commonly classified into tremor-dominant (TD-PD) and postural instability/gait difficulty-dominant (PIGD-PD) motor subtypes based on Movement Disorder Society-Unified Parkinson’s Disease Rating Scale (MDS-UPDRS-III) criteria. These motor phenotypes are associated with distinct clinical trajectories, progression rates, and prognostic profiles, suggesting partially divergent underlying neurobiological mechanisms.^5–6^ TD-PD has been associated with greater involvement of cerebello-thalamo-cortical circuitry, whereas PIGD-PD has been linked to more prominent basal ganglia and fronto-striatal dysfunction.^8–13^

Converging evidence from neuroimaging, neuropathological, and connectivity studies supports these subtype-associated differences in large-scale motor network involvement.^7–12^ Whether these subtype-associated network differences translate into clinically meaningful differences in ICD symptoms remains unknown. Previous studies have been limited by small sample sizes, task-based measures of impulsivity, or narrow operational definitions.^15^ We therefore examined whether clinically measured ICD symptoms differ between TD-PD and PIGD-PD motor subtypes in a large, well-characterized cohort of early PD.

## METHODS

### Study Design, Data Source, and Participants

This study was a cross-sectional observational analysis using data from the Parkinson’s Precision Medicine Initiative (PPMI), an ongoing, multicenter, longitudinal cohort study designed to identify clinical, imaging, and biological markers of PD progression. PPMI enrolls individuals with early PD using standardized clinical assessments and harmonized data collection protocols across participating sites. Data used in the preparation of this article were obtained on 2025-10-02 from the Parkinson’s Precision Medicine Initiative (PPMI) database (https://www.ppmi-info.org/access-data-specimens/download-data), RRID:SCR_006431.^5^ For up-to-date information on the study, visit www.ppmi-info.org. This analysis used data openly available from PPMI.^5^ Participants were included if they had available assessments of ICD symptoms and corresponding motor evaluations required for motor subtype classification.

### TD-PD and PIGD-PD Classification

Motor subtypes were classified as TD-PD or PIGD-PD based on the criteria derived from UPDRS Part II and Part III motor items.^6–9^ Briefly, tremor and PIGD subscores were calculated using standardized groupings of UPDRS items reflecting tremor severity and postural instability/gait difficulty, respectively. These groupings were derived according to previously published motor subtype classification criteria.^6^ A tremor-to-PIGD ratio was then used to classify participants as TD-PD or PIGD-PD according to previously published thresholds.^6^

Motor subtypes were classified using established tremor-to-PIGD ratio criteria derived from the Movement Disorder Society-Unified Parkinson’s Disease Rating Scale (MDS-UPDRS) Part II and Part III assessments. Participants with a tremor/PIGD ratio ≥1.5 were classified as TD-PD, those with a ratio ≤1.0 were classified as PIGD-PD, and those with ratios between 1.0 and 1.5 were considered indeterminate.^6–9^ Participants whose scores did not meet criteria for a definitive TD-PD or PIGD-PD classification were excluded from subtype comparisons.^6–9^

### Impulsivity Assessment

ICD symptoms were assessed using the QUIP-CS, a validated screening instrument designed to capture clinically relevant ICD behaviors in PD.^4^ The QUIP-CS evaluates 13 items across six behavioral domains, including pathological gambling (2 items), compulsive buying (2 items), compulsive sexual behavior (2 items), compulsive eating (2 items), hobbyism/punding/others (3 items), and medication overuse (2 items). Individual items are scored dichotomously (“yes” or “no”), with optional “not applicable” responses for 2 items of “medication overuse”. We used multiple levels of analysis derived from QUIP-CS scoring to characterize ICD symptoms.

### Characterization of ICD Symptoms: Multi-level Analyses

ICD symptoms were evaluated across three levels of analysis designed to capture complementary aspects of impulsive-compulsive symptoms. In the first level of analysis (*Level 1*), the presence of impulsivity on QUIP-CS was defined according to the standard screening design of the instrument, whereby endorsement of any of the 13 QUIP-CS items (“yes” response) was classified as *screening-positive* for ICD symptoms. Participants with no endorsed items were classified as *screening-negative*. This binary categorization reflects the intended screening structure of the QUIP-CS. In the second level of analysis (*Level 2*), ICD symptoms were quantified using a normalized QUIP-CS score calculated as the proportion of endorsed items divided by the total number of scorable items:

#### Normalized QUIP-CS score = (# endorsed items) / (# scorable items)

Normalization accounts for variability in the number of scorable items resulting from optional “not applicable” responses and permits direct comparison of QUIP-CS scores across participants. The normalized score was analyzed both as a continuous variable and categorically. For categorical analyses, a threshold of ≥ 0.25 was used to indicate *higher* ICD; based on the number of scorable items, this threshold corresponded to endorsement of approximately three to four QUIP-CS items. Scores of < 0.25 were categorized as *lower* ICD. Finally, in the third level of analysis (*Level 3*), baseline QUIP-CS assessments were paired with the nearest eligible MDS-UPDRS assessment occurring within a prespecified ± 3-month interval to address potential timing mismatches between behavioral and motor assessments.^19, 20^ Of note, when multiple eligible MDS-UPDRS assessments occurred within this interval, the assessment with the smallest absolute date difference was retained. The categorical and continuous normalized QUIP-CS analyses from Level 2 were also investigated using the temporally aligned dataset. These multi-level analyses were designed to enhance temporal validity while preserving sample size. The overall study design, participant selection, analytic framework, and temporal alignment strategy are illustrated in **Figure 1**.

**Figure 1.**
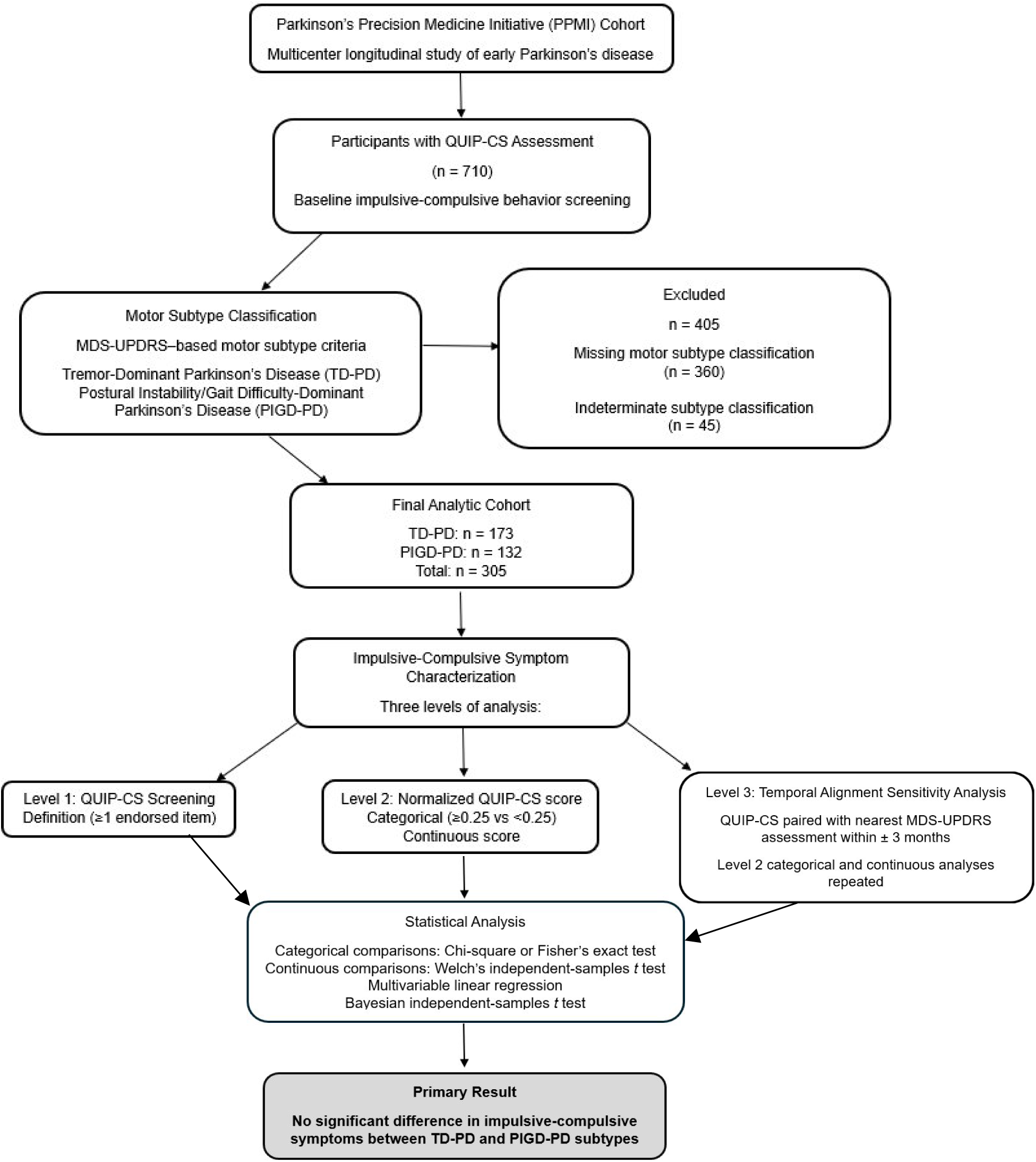
Study Cohort and Analytic Framework. Participants with Questionnaire for Impulsive-Compulsive Disorders in Parkinson’s Disease-Current Short Form (QUIP-CS) assessments (n = 710) from the Parkinson’s Precision Medicine Initiative (PPMI) cohort were classified into Parkinson disease motor subtypes using Movement Disorder Society-Unified Parkinson’s Disease Rating Scale (MDS-UPDRS)–based criteria. Participants with missing motor subtype classification (n = 360) or indeterminate subtype classification (n = 45) were excluded, resulting in a final analytic cohort of 305 participants consisting of the Tremor-Dominant (TD-PD; n = 173) and Postural Instability/Gait Difficulty-Dominant (PIGD-PD; n = 132) motor subtypes. Impulsive-compulsive symptoms were evaluated using three complementary levels of analysis: QUIP-CS screening definition (Level 1); normalized QUIP-CS score analyzed categorically and continuously (Level 2); and a temporal alignment sensitivity analysis (Level 3). For Level 3, QUIP-CS assessments were paired with the nearest MDS-UPDRS assessment within ± 3 months, and both Level 2 analyses were repeated. Statistical comparisons were performed between TD-PD and PIGD-PD groups to assess differences in impulsive-compulsive behaviors.

To evaluate the robustness of the primary findings, prespecified subgroup analyses were performed according to sex (male vs female), age (≥ 62 vs < 62 years), and dopaminergic medication use (on vs not on therapy). A multivariable linear regression model was used to examine the association between motor subtype and normalized QUIP-CS score after adjustment for age, sex, education, disease duration, and baseline levodopa equivalent daily dose (LEDD). LEDD was modeled continuously per 100-mg/day increase with normalized QUIP-CS score specified as the dependent variable.^21^ Bayesian independent-samples *t* testing was performed for the primary normalized-score comparison to quantify evidence supporting either the null or alternative hypothesis using Bayes factors as described by Dienes.^23^

### Statistical Analysis

The continuous normalized QUIP-CS scores were compared between motor subtypes using Welch’s independent-samples *t* tests to account for potential unequal variances. The categorical QUIP-CS outcomes were compared between TD-PD and PIGD-PD motor subtypes using chi-square tests when all expected cell counts were ≥ 5. Fisher’s exact tests were used when expected cell counts were < 5. All statistical tests were two-sided, and statistical significance was defined as a *p* value < 0.05. Statistical analyses were performed using Microsoft Excel (Microsoft Corporation, Redmond, WA, USA). Multivariable linear regression and Bayesian analyses were performed using JASP version 0.98.1 (JASP Team, 2026).^22^ All analyses were performed in the final analytic cohort of 305 participants.

### Data Availability

Deidentified data used in this study are available to qualified investigators through the PPMI database, subject to PPMI data-access and data-use requirements.

## RESULTS

### Cohort Characteristics

Following application of inclusion criteria, motor subtype classification, and assessment availability requirements, a total of 305 participants with PD were included in the final analytic sample. Of these, 173 participants were classified as TD-PD and 132 as PIGD-PD based on UPDRS derived criteria. The mean disease duration of the final analytic cohort was 0.6 years, consistent with an early-stage PD population. Participant selection and analytic cohort derivation are illustrated in **Figure 1**.

Across all levels of analysis, the number of participants classified as TD-PD and PIGD-PD remained consistent. Overall, ICD symptoms at baseline were present in 65 participants (21.3%). Demographic and clinical characteristics of the analytic cohort stratified by motor subtype are summarized in **Table 1**.

**Table 1.** Demographic and Clinical Characteristics of TD-PD and PIGD-PD Participants.

| **Characteristic** | **TD-PD**<br>**(n = 173)** | **PIGD-PD**<br>**(n = 132)** | ***p* Value** |
| --- | --- | --- | --- |
| Age, years, mean (SD) | 62.6 (9.6) | 61.9 (10.0) | 0.560 |
| Male sex, n (%) | 109 (63.0%) | 80 (60.6%) | 0.669 |
| Education, years, mean (SD) | 16.0 (2.9) | 15.1 (3.2) | 0.012 |
| Disease duration, years, mean (SD) | 0.6 (0.6) | 0.5 (0.5) | 0.246 |
| MoCA score, mean (SD) | 26.9 (2.2) | 27.1 (2.4) | 0.454 |
| MDS-UPDRS-III score, mean (SD) | 23.3 (9.8) | 20.3 (10.5) | 0.010 |
| Dopamine agonist use, n (%) | 69 (39.9%) | 43 (32.6%) | 0.191 |

Values are presented as mean (SD) or n (%).
Abbreviations: PD = Parkinson's Disease; MoCA = Montreal Cognitive Assessment; MDS-UPDRS-III = Movement Disorder Society-Unified Parkinson's Disease Rating Scale Part III; TD-PD = Tremor-Dominant; PIGD-PD = postural instability/gait difficulty-dominant.
*P* values were calculated using Welch's *t* tests for age, education, disease duration, MoCA, and MDS-UPDRS-III score, and Pearson's chi-square test for sex and dopamine agonist use.

### Comparison of ICD Symptoms: TD-PD vs PIGD-PD

#### QUIP-CS Screening Definition (positive = any item endorsed, Level 1)

Using the standard screening definition of the QUIP-CS (≥ 1 endorsed item), ICD positivity was observed in 37 of 173 (21.4%) TD-PD participants and 28 of 132 (21.2%) PIGD-PD participants. Our results showed that there was no significant association between motor subtype and QUIP-CS screening positivity (OR = 1.01, 95% CI 0.58-1.76, *p* = 0.970) (**Table 2**).

**Table 2.** Comparison of ICD Symptoms Between TD-PD and PIGD-PD Subtypes.

| Level of Analysis | TD-PD (n = 173) | PIGD-PD (n = 132) | p Value |
| --- | --- | --- | --- |
| Level 1: QUIP-CS screening definition, n (%) | 37/173 (21.4%) | 28/132 (21.2%) | 0.970* |
| Level 2: Normalized QUIP-CS score (Categorical) $\geq 0.25$, n (%) | 6/173 (3.5%) | 6/132 (4.5%) | 0.632* |
| Level 2: Normalized QUIP-CS score (Continuous), mean (SD) | 0.0338 (0.0891) | 0.0384 (0.0973) | 0.674<sup>#</sup> |
| Level 3: Temporally aligned analysis of normalized QUIP-CS score (Categorical) $\geq 0.25$, n (%) | 6/173 (3.5%) | 6/132 (4.5%) | 0.632* |
| Level 3: Temporally aligned analysis of normalized QUIP-CS score (Continuous), mean (SD) | 0.0338 (0.0891) | 0.0384 (0.0973) | 0.674<sup>#</sup> |

Impulsive-compulsive symptoms by motor subtype across three levels of analysis using the Questionnaire for Impulsive-Compulsive Disorders in Parkinson's Disease-Current Short Form (QUIP-CS).
Values are presented as mean (SD) or n (%). \*Pearson's chi-square test; <sup>#</sup>Welch's independent-samples *t* test.
Abbreviations: ICD = Impulse Control Disorder; PD = Parkinson's Disease; TD-PD = Tremor-Dominant; PIGD-PD = postural instability/gait difficulty-dominant.

#### Normalized QUIP-CS Score (Level 2)

We first categorized the normalized QUIP-CS score using the prespecified cut-off (≥ 0.25 vs < 0.25). Scores of ≥ 0.25 were observed in 6 of 173 (3.5%) TD-PD participants and 6 of 132 (4.5%) PIGD-PD participants. There was no significant association between motor subtype and categorical normalized QUIP-CS score (*p* = 0.632) (**Table 2**). We then analyzed the normalized QUIP-CS score as a continuous measure and found that mean scores were similar between the TD-PD and PIGD-PD subtypes, with no significant difference between groups (*p* = 0.674).

#### Analyses by Temporal Alignment (Level 3)

To evaluate the robustness of the findings to potential timing mismatches between behavioral and motor assessments, the normalized QUIP-CS analyses were repeated after pairing baseline QUIP-CS assessments with the nearest eligible MDS-UPDRS assessment occurring within ± 3 months. The temporally aligned dataset included the same 305 participants. Results remained unchanged for both the categorical and continuous normalized scoring approaches (Table 2).

### Effects of sex, age, and dopaminergic medication

Considering that sex, age, and dopaminergic medication use can contribute to ICD symptoms, the effects of these factors were further examined. Stratified analyses by sex, age, and dopaminergic medication use demonstrated consistent findings across all levels of analysis. In the overall cohort, ICD symptoms were present in 65 participants (21.3%). The proportion of participants with ICD symptoms was similar across subgroups: male (37/189, 19.6%) vs female (28/116, 24.1%); age ≥ 62 years (32/165, 19.4%) vs < 62 years (33/134, 24.6%); and dopaminergic medication use (+) (24/112, 21.4%) vs (–) (41/193, 21.2%). Using binary classification (Level 1), no meaningful differences in ICD symptoms were observed between TD-PD and PIGD-PD subtypes within any subgroup. Using normalized scoring (Level 2), the normalized QUIP-CS scores remained low and similarly distributed across subgroups, with no consistent differences between motor subtypes. Overall, findings were consistent across all levels of analysis, supporting the absence of a significant association between motor subtype and ICD symptoms.

A multivariable linear regression model demonstrated that motor subtype remained unassociated with normalized QUIP-CS score after adjustment for baseline levodopa equivalent daily dose (LEDD), age, sex, education, and disease duration (*β* = −0.0063, 95% CI −0.0292 to 0.0167, *p* = 0.590).

Bayesian independent-samples *t* testing of normalized QUIP-CS scores provided moderate evidence supporting the null hypothesis of no difference between TD-PD and PIGD-PD participants (BF_01_ = 7.21). The observed data were approximately 7.2 times more likely under the null hypothesis than under the alternative hypothesis.

## DISCUSSION

In this cross-sectional analysis of a well-characterized early PD cohort, ICD symptoms did not differ between TD-PD and PIGD-PD motor subtypes. Across multiple levels of analysis, including the standard screening definition, normalized scoring, and temporally aligned analyses, no significant association between motor subtype and ICD symptoms was observed. These findings support the absence of a clinically meaningful association between motor subtype and ICD symptoms. Taken together, these findings suggest that ICDs are largely independent of PD motor phenotype. This pattern remained consistent after adjustment for baseline LEDD and other clinical covariates and was further supported by Bayesian analysis, providing evidence in favor of the null hypothesis model.

Previous studies examining the relationship between PD motor subtype and impulsivity have yielded inconsistent findings, often due to small sample sizes, task-based measures of motor impulsivity, or narrow operational definitions. The most direct prior investigation reported greater susceptibility to motor impulsivity in PIGD-PD compared with TD-PD based on laboratory response-inhibition tasks but did not evaluate clinically relevant ICD behaviors or broader impulsive-compulsive symptom domains.^17^ The present study extends this literature by applying prespecified multi-level analyses of ICD symptoms in a large, multicenter cohort with standardized motor and behavioral assessments. By demonstrating consistent null findings across multiple levels of analysis and temporal alignment restrictions, the current results help clarify previously inconclusive evidence and suggest that ICDs do not meaningfully differ between TD-PD and PIGD-PD motor subtypes in early PD.^1–3,8–9^

Although the cerebellum has been increasingly implicated in non-motor functions such as timing, prediction, and behavioral regulation, evidence linking cerebellar motor circuitry to clinically relevant impulsive-compulsive behaviors in PD remains limited.^16^ Prior structural, functional, and neuropathologic studies have suggested that TD-PD and PIGD-PD differ in large-scale motor network involvement, with relatively greater cerebello-thalamo-cortical involvement in TD-PD and more prominent frontostriatal and basal ganglia-related abnormalities in PIGD-PD.^8–13^ Despite these subtype-related circuit differences, we did not find corresponding differences in QUIP-CS-defined ICD symptoms. These findings are consistent with previous work implicating basal ganglia and frontostriatal circuits in ICDs rather than suggesting a dominant contribution from motor subtype-specific circuitry. Neuroimaging and neurophysiological studies have demonstrated associations between impulsive-compulsive behaviors and alterations in affective and sensorimotor striatal circuits, frontostriatal connectivity, and reward-related processing, largely independent of motor phenotype.^15–16,18^

Strengths of this study include the use of a large, well-characterized multicenter cohort, standardized assessments of motor phenotype and ICD symptoms, and multiple levels of analysis designed to address potential sources of misclassification and timing bias. The application of temporal alignment criteria further supports the robustness of the findings.^5,8^ Our present study has several limitations. First, ICD symptoms were assessed using the QUIP-CS, a validated screening instrument rather than a diagnostic interview, which is inherent to the original design of PPMI and may limit sensitivity for identifying formally diagnosed ICDs.^4^ Second, the cross-sectional design precludes assessment of longitudinal changes in ICD symptoms or potential evolution of motor subtype over time. Furthermore, motor subtype classification represents a clinical construct that may evolve with disease progression, and subtype stability over time was not assessed.^8,14^ Third, the proportion of participants with normalized QUIP-CS scores of ≥ 0.25 was low in this early PD cohort, which may limit power to detect rare subtype-specific effects. Future studies should evaluate ICD symptoms in later disease stages, when exposure to dopamine agonists is more common and may exert a stronger influence on impulsive-compulsive behaviors.

## CONCLUSION

In this null-hypothesis analysis of early PD, clinically measured ICD symptoms were largely independent of Parkinson disease motor phenotype. These findings suggest that differences in motor phenotype-associated neural circuitry do not translate into clinically meaningful differences in ICDs, providing a clearer framework for future mechanistic and longitudinal studies of ICDs in PD.

## Supporting information

PPMI Author List June 2025

## Data Availability

Deidentified data used in this study are available to qualified investigators through the Parkinson's Precision Medicine Initiative (PPMI) database, subject to PPMI data-access and data-use requirements.

https://www.ppmi-info.org/access-data-specimens/download-data

https://doi.org/10.5281/zenodo.21630900

## Acknowledgement

The README, data preparation workflow, and statistical analysis workflow supporting this study are publicly available on Zenodo [https://doi.org/10.5281/zenodo.21630900]. PPMI – a public-private partnership – is funded by the Michael J. Fox Foundation for Parkinson’s Research and funding partners, including AbbVie, Alamar Biosciences, Aligning Science Across Parkinson’s (ASAP), Arrowhead Pharma, Arvinas, AskBio, BIAL, BioArctic, Biohaven, BlueRock Therapeutics, Bristol Myers Squibb, Calico Labs, Capsida Biotherapeutics, Critical Path Institute, DaCapo Brainscience, Denali, Edmond J. Safra Foundation, Eli Lilly, Gain Therapeutics, GE Healthcare, Genentech, GSK, Insitro, Johnson & Johnson Innovative Medicine, Lundbeck, Merck, Neumora, Neuron23, Novartis, Olink, Regeneron, Roche, Sanofi, Tenvie, UCB, Vanqua Bio, Voyager Therapeutics and The Weston Family Foundation.

## Author Contributions

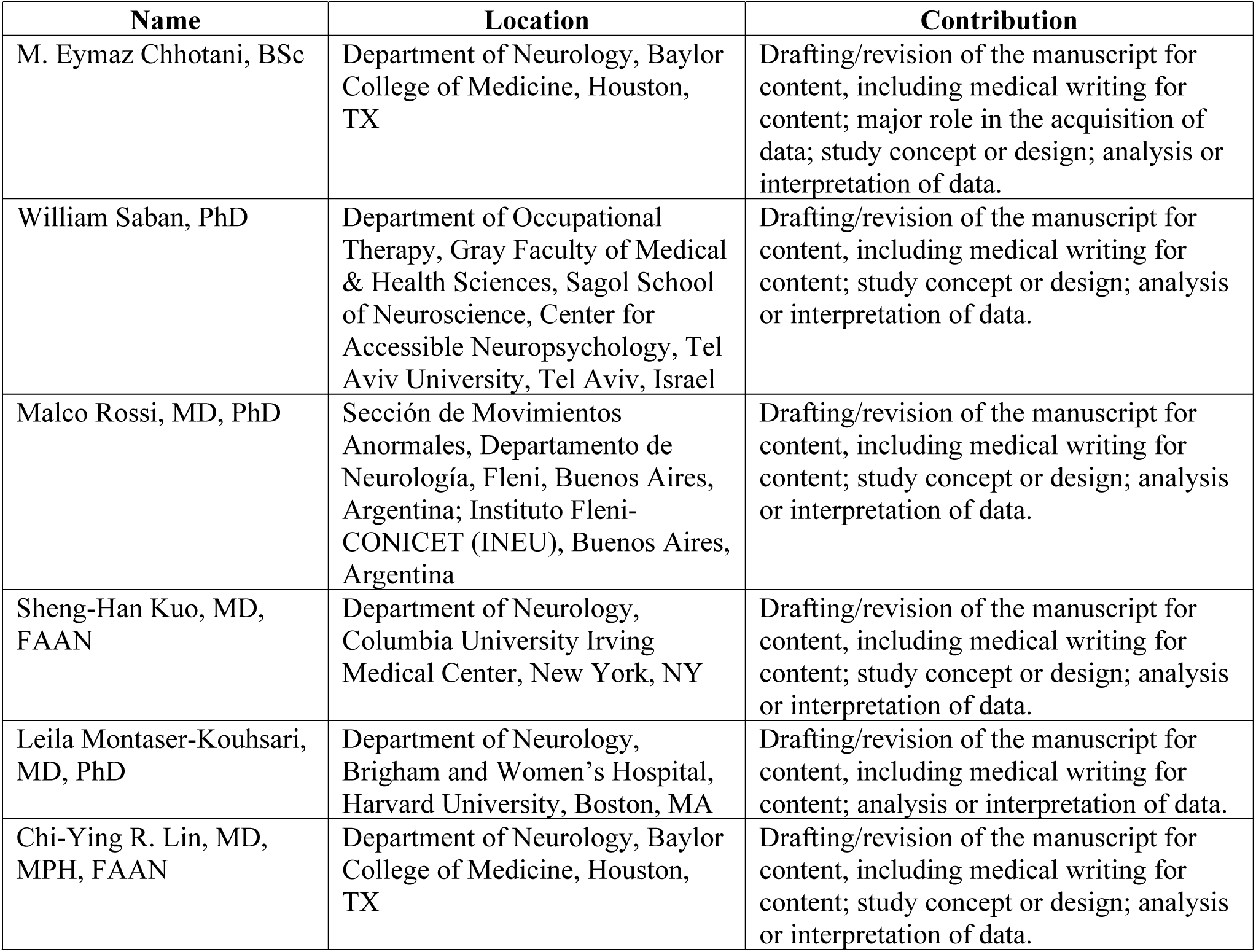

## Supplemental Material

The PPMI Study Committees, Cores, and Collaborators Author List (June 2025) is provided as supplemental material.

