## Supplementary material for "Impulse Control Disorders Are Independent of Parkinson Disease Motor Subtypes": PPMI Author List June 2025

Parkinson's Progression Markers Initiative

### PPMI STUDY COMMITTEES, CORES, AND COLLABORATORS

#### PPMI EXECUTIVE STEERING COMMITTEE

Kenneth Marek, MD[1] (Principal Investigator); Tanya Simuni, MD[2]; Andrew Siderowf, MD[3]; Caroline Tanner, MD[4]; Thomas F Tropea, DO[1]; Tatiana Foroud, PhD[5]; Lana Chahine, MD[6]; Brit Mollenhauer, MD[7]; Kalpana Merchant, MD[2]; Douglas Galasko, MD[8]; Christopher Coffey, PhD[9]; Kathleen Poston, MD[10]; Roseanne Dobkin, PhD[11]; Ethan Brown, MD[4]; Roy Alcalay, MD[12]; Dan Weintraub, MD[3]; Emily Flagg, BA[1]; Kimberly Fabrizio, BA[1]

#### PPMI STEERING COMMITTEE

Susan Bressman, MD[13]; Cornelis Blauwendraat, PhD[14]; Paola Casalin, PhD[15]; Sonya Dumanis, PhD[14]; Raymond James, RN[16]; Karl Kieburtz, MD[17]; Sneha Mantri, MS[18]; Werner Poewe, MD[19]; Michael Schwarzschild, MD[20]; John Seibyl[1], MD; David Standaert, PhD[21]; Duygu Tosun-Turgut, PhD[4]

#### MICHAEL J. FOX FOUNDATION

Sohini Chowdhury, MA[22]; Jamie Eberling, PhD[22]; Mark Frasier, PhD[22]; Leslie Kirsch, EdD[22]; Katie Kopil, PhD[22]; Maggie Kuhl, BA[22]; Alyssa O'Grady, BA[22]; Todd Sherer, PhD[22]; Tawny Willson, MBS[22]

#### PPMI STUDY CORES

*Project Management Core:* Emily Flagg, BA[1]
*Site Management Core:* Tanya Simuni, MD[2]; Bridget McMahon, BS[1]
*Data Strategy and Technical Operations:* Craig Stanley, PhD[1]; Kim Fabrizio, BA[1]
*Data Management Core:* Dixie Ecklund, MBA[9], MSN; Christine Kohnen, PhD[9]
*Screening Core:* Tatiana Foroud, PhD[5]; Laura Heathers, BA[5]; Christopher Hobbick, BSCE[5]; Gena Antonopoulos, BSN[5]
*Imaging Core:* John Seibyl, MD[1]; Kathleen Poston, MD[10]
*Statistics Core:* Christopher Coffey, PhD[9]; Chelsea Caspell-Garcia, MS[9]; Michael Brumm, MS[9]
*Bioinformatics Core:* Arthur Toga, PhD[23]; Karen Crawford, MLIS[23]
*Biorepository Core:* Tatiana Foroud, PhD[5]; Jan Hamer, BS[5]
*Biologics Review Committee:* Brit Mollenhauer, MD[7]; Doug Galasko, MD[8]; Kalpana Merchant, MD[2]
*Genetics Core:* Andrew Singleton, PhD[24]
*Pathology Core:* Tatiana Foroud, PhD[5]; Dirk Keene, MD[5]
*Found:* Caroline Tanner, MD[4]; Ethan Brown, MD[4]
*PPMI Online:* Carlie Tanner, MD[4]; Ethan Brown, MD[4]; Lana Chahine, MD[6]; Roseann Dobkin, PhD[11]; Monica Korell, MPH[4]

#### PPMI SITE INVESTIGATORS

Neha Prakash MD[1]; Tanya Simuni, MD[2]; Nabila Dahodwala MD[3]; Caroline Tanner, MD[4]; Lana Chahine MD[6]; Brit Mollenhauer MD[7]; Sebastian Schade MD[7]; Douglas Galasko, MD[8]; Anat Mirelman PhD[12]; Roy Alcalay MD[12];Katherine Leaver MD[13]; Marie Saint-Hilaire MD[16]; Ruth Schneider MD[17]; Christopher Tarolli MD[17]; Werner Poewe, MD[19]; Aleksandar Videnovic MD[20]; David Standaert PhD[21]; Marissa Dean, MD[21]; Sonja Jonsdottir PhD[25]; Rejko Krueger MD[25]; Claire Pauly PhD[25]; Stewart Factor DO[26]; Penelope Hogarth MD[26]; Robert Hauser MD[28]; Amy Amara PhD[29]; Michelle Fullard MD[29]; Cyrus Zabetian MD[30]; Hubert Fernandez MD[31]; Kathrin Brockmann MD[32]; Isabel Wurster PhD[32]; Yen Tai PhD[33]; Paolo Barone PhD[34]; Marina Picillo MD[34]; Stuart Isaacson MD[35]; Alberto Espay MD[36]; Eduardo Tolosa PhD[37]; Javier Ruiz Martinez PhD[38]; Leonidas Stefanis PhD[39]; Kelvin Chou MD[40]; Lorraine Kalia MD[41]; Connie Marras PhD[41]; David Grimes MD[42]; Tiago Mestre PhD[42]; Rajesh Pahwa MD[43]; Mark Lew MD[44]; Holly Shill MD[45]; Shyamal Mehta MD[46]; Giulietta Riboldi MD[47]; Nikolaus McFarland PhD[48]; Ron Postuma MD[49]; Zoltan Mari MD[50]; David Ledingham MD[51]; Nicola Pavese PhD[51]; Michele Hu PhD[52]; Norbert Brueggemann MD[53], ; Christine Klein MD[53]; Bastiaan Bloem PhD[54]; Cristina Simonet PhD[55]; Alastair Noyce PhD[55]; Anette Janzen PhD[56]; David Pedrosa MD[56]; Wolfgang Oertel PhD[56]; Njideka Okubadejo MD[57] David Shprecher DO[58]; Arjun Tarakad MD[59]; Emile Moukheiber MD[60]

#### PPMI SITE COORDINATORS

Joy Antala[1]; Carla Aranda[2]; Karen Williams[2]; Sophia Melton[2]; Karina Benson[2]; Ashwini Ramachandran[3]; Danielle Potts[3]; Grace LaMoure[3]; Ritikha Vengadesh[3]; Ryan Manzler[3]; Jaime Heller[4]; Primi Ranola[4]; Farah Kausar[4]; Sherri Mosovsky[6]; Diana Willeke[7]; Elizabeth Kalinkara-Gomez[7]; Janelle Rodriguez[8]; Nobuko Kemmotsu[8]; May Eshel[12]; Deborah Raymond[13]; Abigail Desrosiers[16]; Raymond James[16]; Lauren Jackson[17]; Iris Egner[19]; Wesley Schlett[20]; Courtney Blair[21]; Lauren Ruffrage[21]; Berenice Sevilla[25]; Barbara Sommerfeld[26]; Dustin Le[27]; Erica Botting[28]; Gabriella Mazur[28]; Daniele Derlein[29]; Evan Doll[29]; Ying Liu[29]; Ciera Cobb[30]; Olivia Masiewicz[30]; Jennifer Mule[31]; Michael Morsillo[31]; Ella Hilt[32]; Aldazier Jakiran[33]; Dominga Valentino[34]; Lisbeth Pennente[35]; Bobbie Stubbeman[36]; Alicia Garrido[37]; Valeria Ravasi[37]; Ioana Croitoru[38]; Christos Koros[39]; Nikolas Papagiannakis[39]; Frank Ferrari[40]; Mengyu Zheng[41]; Shawna Reddie[42]; Alicia Alejandra[43]; Andrea Gray[43]; Alejandra Valenzuela[44]; Caitlin Goodman [45]; Sara Dresler[46]; Neil Santos[46]; Fahrial Esha[47]; Kyle Rizer[48]; Nadine Zablith[49]; Liliana Dumitrescu[50]; Debra Galley[51]; Victoria Kate Foster[51]; Jamil Razzaque[52]; Madita Grümmer[53]; Yara Krasowski[54]; Natalie Donkor[55]; Elisabeth Sittig[56]; Oluwadamilola Ojo[57]; Kelly Clark[58]; Rory Mahabir[59]; Kori Ribb[60]; Shamera Willoughby[60]

#### INSTITUTIONS AND AFFILIATIONS


1. Institute for Neurodegenerative Disorders; New Haven, CT, USA
2. Northwestern University; Evanston, IL, USA
3. University of Pennsylvania; Philadelphia, PA, USA
4. University of California, San Francisco; San Francisco, CA, USA
5. Indiana University; Indianapolis, IN, USA
6. University of Pittsburgh; Pittsburgh, PA, USA
7. Paracelsus-Elena Klinik; Kassel, Germany
8. University of California, San Diego, San Diego, CA, USA
9. University of Iowa; Iowa City, IA, USA
10. Stanford University; Stanford, CA, USA
11. Rutgers University; New Brunswick, NJ, USA
12. Tel Aviv Sourasky Medical Center; Tel Aviv, Israel
13. Mount Sinai Beth Israel; New York, NY, USA
14. Coalition for Aligning Science; Chevy Chase, MD, USA
15. BioRep; Milan, Italy
16. Boston University School of Medicine; Boston, MA, USA
17. University of Rochester; Rochester, NY, USA
18. Duke University; Durham, NC, USA
19. University of Innsbruck; Innsbruck, Austria
20. Massachusetts General Hospital; Boston, MA, USA
21. University of Alabama at Birmingham; Birmingham, AL, USA
22. The Michael J. Fox Foundation for Parkinson's Research; New York, NY, USA
23. Laboratory of Neuroimaging (LONI), USC; Los Angeles, CA, USA
24. National Institute on Aging, NIH; Bethesda, MD, USA
25. University of Luxembourg; Esch-sur-Alzette, Luxembourg
26. Emory University; Atlanta, GA, USA
27. Oregon Health and Science University; Portland, OR, USA
28. University of South Florida; Tampa, FL, USA
29. University of Colorado; Aurora, CO, USA
30. VA Puget Sound Health System; Seattle, WA, USA
31. Cleveland Clinic; Cleveland, OH, USA
32. University of Tubingen; Tubingen, Germany
33. Imperial College of London; London, UK
34. University of Salerno; Salerno, Italy
35. Parkinson's Disease and Movement Disorders Center; Boca Raton, FL, USA
36. University of Cincinnati; Cincinnati, OH, USA
37. Hospital Clinic of Barcelona; Barcelona, Spain
38. Hospital Universitario Donostia; San Sebastian, Spain
39. University of Athens; Athens, Greece
40. University of Michigan; Ann Arbor, MI, USA
41. Toronto Western Hospital; Toronto, Canada
42. The Ottawa Hospital; Ottawa, Canada
43. University of Kansas Medical Center; Kansas City, KS, USA
44. Keck School of Medicine of the University of Southern California; Los Angeles, CA, USA
45. Barrow Neurological Institute; Phoenix, AZ, USA
46. Mayo Clinic Arizona; Scottsdale, AZ, USA
47. NYU Langone Medical Center; New York, NY, USA
48. University of Florida; Gainesville, FL, USA
49. Montreal Neurological Institute and Hospital/McGill; Montreal, QC, Canada
50. Cleveland Clinic-Las Vegas Lou Ruvo Center for Brain Health; Las Vegas, NV, USA
51. Clinical Ageing Research Unit; Newcastle, UK
52. John Radcliffe Hospital Oxford and Oxford University; Oxford, UK
53. University of Luebeck; Luebeck, Germany
54. Radboud University; Nijmegen, Netherlands
55. Queen Mary University of London; London, UK
56. Philipps-University Marburg; Marburg, Germany
57. University of Lagos; Lagos, Nigeria
58. Banner Sun Health Research Institute; Sun City, AZ, USA
59. Baylor College of Medicine; Houston, TX, USA
60. Johns Hopkins University; Baltimore, MD, USA


#### PPMI STUDY FUNDING PARTNER STATEMENT


PPMI – a public-private partnership – is funded by the Michael J. Fox Foundation for Parkinson's Research and funding partners, including 4D Pharma, Abbvie, AcureX, Allergan, Amathus Therapeutics, Aligning Science Across Parkinson's, AskBio, Avid Radiopharmaceuticals, BIAL, BioArctic, Biogen, Biohaven, BioLegend, BlueRock Therapeutics, Bristol-Myers Squibb, Calico Labs, Capsida Biotherapeutics, Celgene, Cerevel Therapeutics, Coave Therapeutics, DaCapo Brainscience, Denali, Edmond J. Safra Foundation, Eli Lilly, Gain Therapeutics, GE HealthCare, Genentech, GSK, Golub Capital, Handl Therapeutics, Insitro, Jazz Pharmaceuticals, Johnson & Johnson Innovative Medicine, Lundbeck, Merck, Meso Scale Discovery, Mission Therapeutics, Neurocrine Biosciences, Neuron23, Neuropore, Pfizer, Piramal, Prevail Therapeutics, Roche, Sanofi, Servier, Sun Pharma Advanced Research Company, Takeda, Teva, UCB, Vanqua Bio, Verily, Voyager Therapeutics, the Weston Family Foundation and Yumanity Therapeutics.
